# Blood biomarkers-defined subgroups show heterogeneity in post-acute COVID-19 syndrome: a rationale for precision medicine

**DOI:** 10.1101/2023.03.31.23288003

**Authors:** Benjamin Charvet, Alexandre Lucas, Justine Pierquin, Joanna Brunel, Steven Fried, Corinne Bernis, Zachary S. Orban, Millenia Jimenez, Barbara A. Hanson, Lavanya Visvabharathy, Igor J. Koralnik, Hervé Perron

## Abstract

Acute COVID-19 can cause a post-infectious syndrome in a significant percentage of patients, with multifacted and long lasting symptoms. We hypothesized that this Post-Acute COVID syndrome (PASC) could result from various underlying causes, which may compromise the demonstration of efficacy for treatments evaluated on cohorts of heterogeneous patients.

To assess the feasibility of stratifying or characterizing subgroups of post-COVID-19 patients consistent with different indications in a precision medicine perspective, we tested serum biomarkers in a pilot cross-sectional study of patients with neuro-cognitive symptoms from the Northwestern University post-COVID-19 clinic (Chicago,USA). Patient health status was evaluated with the use of standardized PROMIS questionnaires and underwent validated cognitive tests with the NIH Toolbox. Serum biomarkers were chosen as proteins known to be involved in the pathogenic features of a neuro-inflammatory disease, i.e., multiple sclerosis, with a final selection of the most discriminant ones. A multi-isotypes serology against SARS-CoV-2 spike and nucleocapsid antigens was performed to allow detailed analyses of the humoral immune status.

Despite the limited numbers of this feasibility study, results showed that clinical data could not differentiate PASC patients with persisting neuro-cognitive impairment, while three major PASC subgroups were identified with serum biomarkers according to the presence or absence of the HERV-W ENV soluble protein combined with neurofilaments light chains and, to a lesser extent, with elevated levels of IL-6. SARS-CoV-2 serological results in PASC compared to healthy controls also revealed a significant increase of anti-Spike and/or Nucleocapsid IgM, IgA and, unexpectedly, IgE. For IgG, a significant difference was observed with Nucleocapsid only since anti-Spike IgG titers were normally elevated in vaccinated controls. This multi-Ig isotypes serology may provide additional information on the infectious and immunological status of individual patients and should be considered in face of a potential viral persistence in some individuals.

Altogether the results show the feasibility of using serum biomarkers to discriminate relevant subgroups or individual patients for precision medicine indications in post-COVID syndromes. This pilot study paves the way to further exploring biological assays for the definition of subtypes of PASC, also called long COVID, useful for the choice of relevant therapeutic strategies.

## 1 Introduction

The COVID-19 pandemic is now known to generate a post-viral syndrome with multifaceted and long-lasting symptoms, variably named “Long COVID”, “Post-COVID” or “Post-Acute sequelae of COVID-19 (PASC). Though the last denomination may suggest lingering post-infectious symptoms, e.g., ischemic tissue lesions following thrombotic events ^1^, many symptoms appear to result from ongoing pathogenic processes with possible onset of chronicity ^2,3^.

In June 2022, the center for disease control (CDC) of the USA published that “more than 40% of adults in the United States reported having COVID-19 in the past, and nearly one in five of those (19%) are currently still having symptoms of long COVID”^4^. In September 2022, the World Health Organization (WHO) published that “at least 17 million people in the WHO European Region experienced long COVID in the first two years of the pandemic; millions may have to live with it for years to come” ^5^. The data points to a secondary pandemic caused by COVID-19, in the form of a post-COVID syndrome of unknown duration and evolution with overlapping symptoms that may result from various pathogenic factors and pathways ^6^. In particular, persisting neurological and cognitive symptoms appear common and affect the quality of life on the long term ^7,8^.

Diagnosing and managing such a complex syndrome is a challenge that requires understanding how to identify potentially differing mechanisms underlying the overlapping symptoms observed in these patients ^9^. There is a need to identify markers which will differentiate between heterogeneous long-COVID subgroups and guide precision medicine where possible. For this purpose, we have selected biomarkers from factors already known to be involved in COVID-19 and also suspected to contribute to certain aspects of long COVID or PACS. We also focused on assays suitable for routine laboratory analyses on serum samples, which can be easily obtained from patients.

In the present pilot study on patients from a post-COVID clinic presenting with persistent symptoms such as abnormal fatigue, sleep disorders and dominant neuro-cognitive symptoms, we have thus studied (i) immunoglobulin isotypes of the humoral response against SARS-CoV-2 spike and nucleocapsid antigens, (ii) the presence of an endogenous retroviral protein, HERV-W ENV, shown to be pathogenic for the immune and nervous systems ^10-12^, with virus-induced expression ^13-15^ and recently shown to predict disease severity when detected on lymphocytes from COVID-19 patients ^16^, (iii) the levels of major cytokines involved in COVID-19 inflammatory responses ^17^ and, because of neuro-cognitive symptoms, (iv) the levels of neurofilament light chain (NfL) known to be accurate biomarkers of neuronal impairment even when quantified in the peripheral blood ^18^.

## 2 Methods

### Participants and clinical assessment

This cross-sectional study was approved by the Northwestern University Institutional Review Board (STU00215866). Blood collection from patients and sample processing followed standard operating procedures with the appropriate approval of the Ethics and Scientific Committees (num. 20210604/04/02). Written informed consent was obtained from all study participants. Importation of the human samples received the authorization IE-2022-2309 from the French ministry. Clinical and demographic data from PASC patients are presented in **Table 1**. PASC patient health status was evaluated with the use of standardized PROMIS questionnaires and underwent validated cognitive tests with the NIH Toolbox ^8,19,20^.

**TABLE 1.**
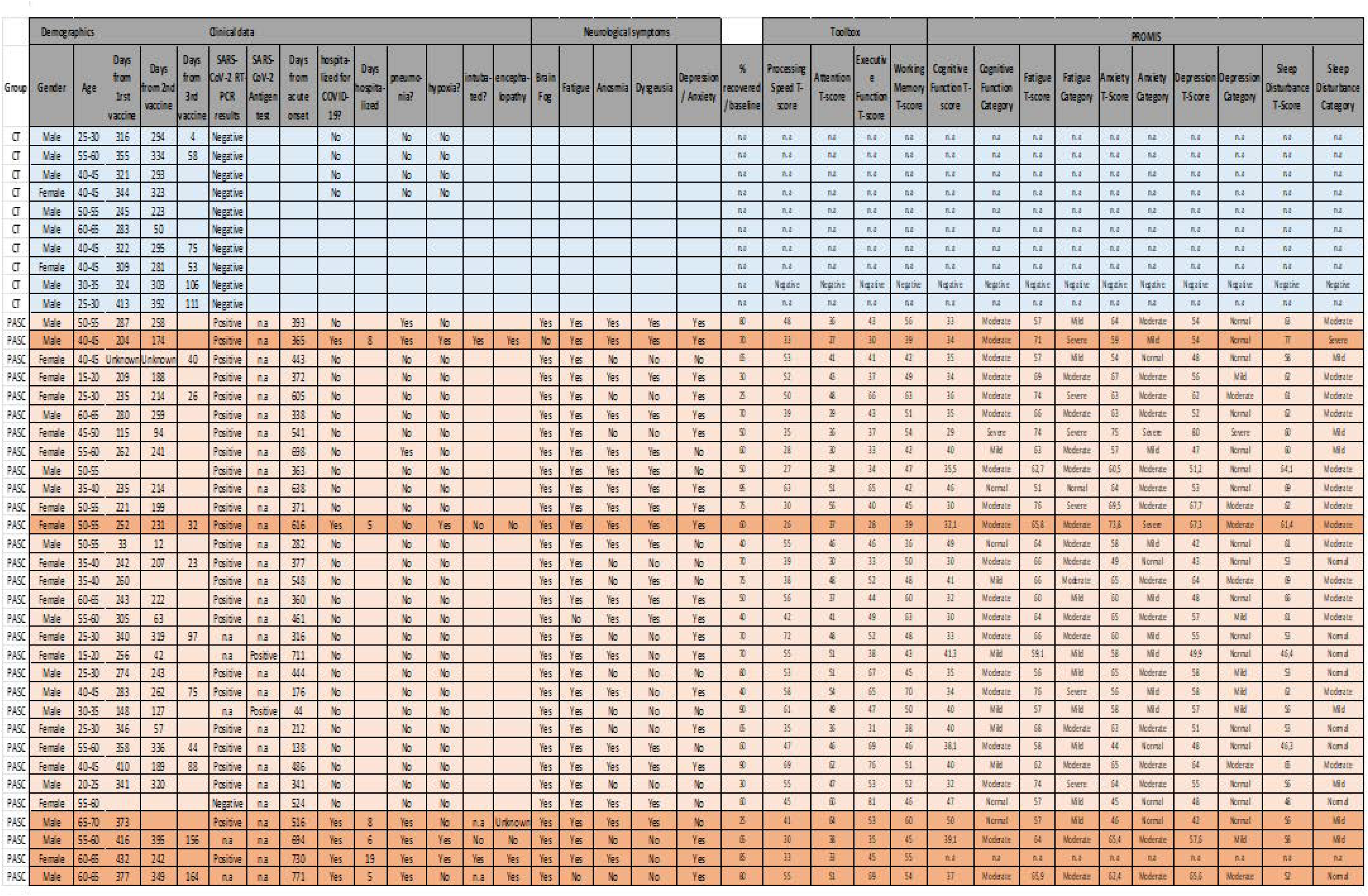

### Immunooassays

The detection of SARS-CoV-2 immunoglobulin isotypes (IgG, IgM, IgA and IgE) were performed on serum by Simple Western technology (ProteinSimple/BioTechne, CA, USA) with manufacturer’s reagents according to their manual. HIS-tagged recombinant proteins of SARS-CoV-2 ((S)Spike, S1 Subunit, S2 Subunit, S1 RBD and Nucleocapsid were used to detect antibody responses to SARS-CoV-2. The area under the curve (AUC) for the electrophoregram peak corresponding to the immunodetection of each protein at the expected size was measured using Compass Software.

For the detection of HERV-W ENV antigen in serum analyses were also performed with simple western-Jess, according to the conditions provided in the patent “Method for the detection of the soluble hydrophilic oligomeric form of HERV-W envelope protein”, published under ref. WO2019201908, with previously described sample treatment (32). The specific signal was expressed as the signal to noise (S/N) ratio, where the noise represents the mean+2SD of the background signals yielded by a panel of sera from healthy blood donors (EFS, Lyon-France). PASC samples were stored in freezers at -80ºC for less than 6 months. All samples were kept frozen after their initial freezing until use (no freeze/thaw cycle before the immunoassay).

Serum levels of NfL (# SPCKB-PS-002448) and IL-6 (# SPCKC-PS-006520) were quantified with the fully automated immunoassay platform, Ella (ProteinSimple/Bio-techne, CA, USA). The lower limits of quantification (LLOQ) were provided by the supplier as a threshold for normal baselines 3 pg/mL for IL-6. Age-standardized Z-scores were calculated for NfLs{Harp, 2022 #18599}. Serum samples were run in triplicate and results in pg/mL were calculated by the instrument software (SimplePlex Explorer, ProteinSimple/Bio-techne, USA).

### RT-qPCR

Total RNA was obtained from PAXgene tubes used to collect whole blood samples and stored at - 20°C. RNA extraction used NucleoSpin RNA Mini Kit (Macherey Nagel, 740955) according to the manufacturer’s protocol. 200 ng of DNase-treated RNA was reverse-transcribed into cDNA using iScript cDNA Synthesis Kit (Bio-Rad, 1708891) according to the manufacturer’s protocol. A control with no-RT was used to confirm the absence of contaminating DNA. 5 ng of initial RNA in RT reaction was used in RT-qPCR for HERV-W *ENV*, HERV-W *POL*, HERV-K *ENV*, SARS-CoV-2 N genes. Primer sequences (5’ to 3’): HERV-K ENV Forward CTGAGGCAATTGCAGGAGTT, Reverse GCTGTCTCTTCGGAGCTGTT; HERV-W ENV Forward GTATGTCTGATGGGGGTGGAG, Reverse CTAGTCCTTTGTAGGGGCTAGAG; HERV-W POL Forward CCTGTACGTCCTGACTCTC, Reverse CTTGGGCTAATGCCTGGCC; SARS-CoV-2 N Forward AAACATTCCCACCAACAG, Reverse CACTGCTCATGGATTGTT; B2M Forward TTACTCACGTCATTCAGCAG, Reverse GATGGATGAAACCCAGACAC. Assays used StepOnePlus instrument (Applied Biosystems) with Platinum SYBR Green (Invitrogen, 11744-500). Housekeeping gene beta-2 microglobulin (*B2M*) normalized the results. A melting curve analysis confirmed the specificity of amplification and the lack of non-specific products. Quantification used the threshold cycle (Ct) comparative method: the relative expression was calculated as follow: 2^-[DCt (sample) - DCt (calibrator)]^ = 2^-DDCt^, where DCt (sample) = [Ct (target gene) – Ct (housekeeping gene)] and the DCt (calibrator) was the mean of DCt of healthy controls.

### Statistics

Statistical analyses were performed using Prism (version 9.0; GraphPad Software, La Jolla, California). Continuous data are expressed as means□±□SD, as indicated. Normal data distribution was assessed using the Shapiro-Wilk or D’Agostino & Pearson’s test, assuming p-values□>□0.05. Statistical differences were determined using Student’s test (two tailed) for compared groups with normal distributions, with Welch’s correction when SDs of samples were significantly different (F test p<0.01) - Mann-Whitney U test (two tailed) was used for compared groups without normal distribution and Wilcoxon signed rank test (two tailed) was used for comparison when a group only or mostly comprised null or very low values for a given threshold. Chi-square (two tailed) test was used for comparison of numbers in categories, replaced by Fischer’s exact test when one category had zero count. Correlation analyses were performed with Spearman test (two tailed). Statistical significance was set at p<0.05.

## 3 Results

A total of 41 subjects (Table 1) were evaluated in this study. Controls (Ctrl) were caregivers or volunteers from hospital (n=10) and PASC patients previously diagnosed with acute COVID-19 and presenting with neuro-cognitive symptoms, (Neuro-PASC; n=31). These included 6 post-hospitalization Neuro-PASC (PNP) and 25 non-hospitalized Neuro-PASC (NNP) patients seen at Northwestern Medicine Neuro-COVID-19 clinic.

### HERV-W ENV detection

Following reports of HERV-W envelope RNA and protein in acute COVID-19 patients correlating with disease severity and markers of T-cell exhaustion ^16^, this study analyzed HERV-W *ENV* RNA from whole blood and levels of soluble envelope protein (W-ENV) in serum from PASC compared to Ctrl. As shown in Figure 1 A-D, about one-half (11/23 with QC-passed RNA) of PASC individuals had elevated RNA levels and about one-third (10/31) had detectable W-ENV protein in serum, which was significantly different from Ctrl for both analyses (Wilcoxon Signed Rank test: Ctrl W=1.00, PASC W=177.00, N=7+23, p=0.0055 for whole blood RNA, and Ctrl W=10.00, PASC W=265.00, N=10+31, p=0.0020 for serum protein). RT-qPCR also detects non-coding RNA and it is known that HERV RNA could play physiological roles as regulatory non-coding long or small RNA ^21,22^. Nevertheless, the presence of circulating W-ENV protein, a potent TLR4 agonist, cannot be neutral^12,23-26^.

**Figure 1.**
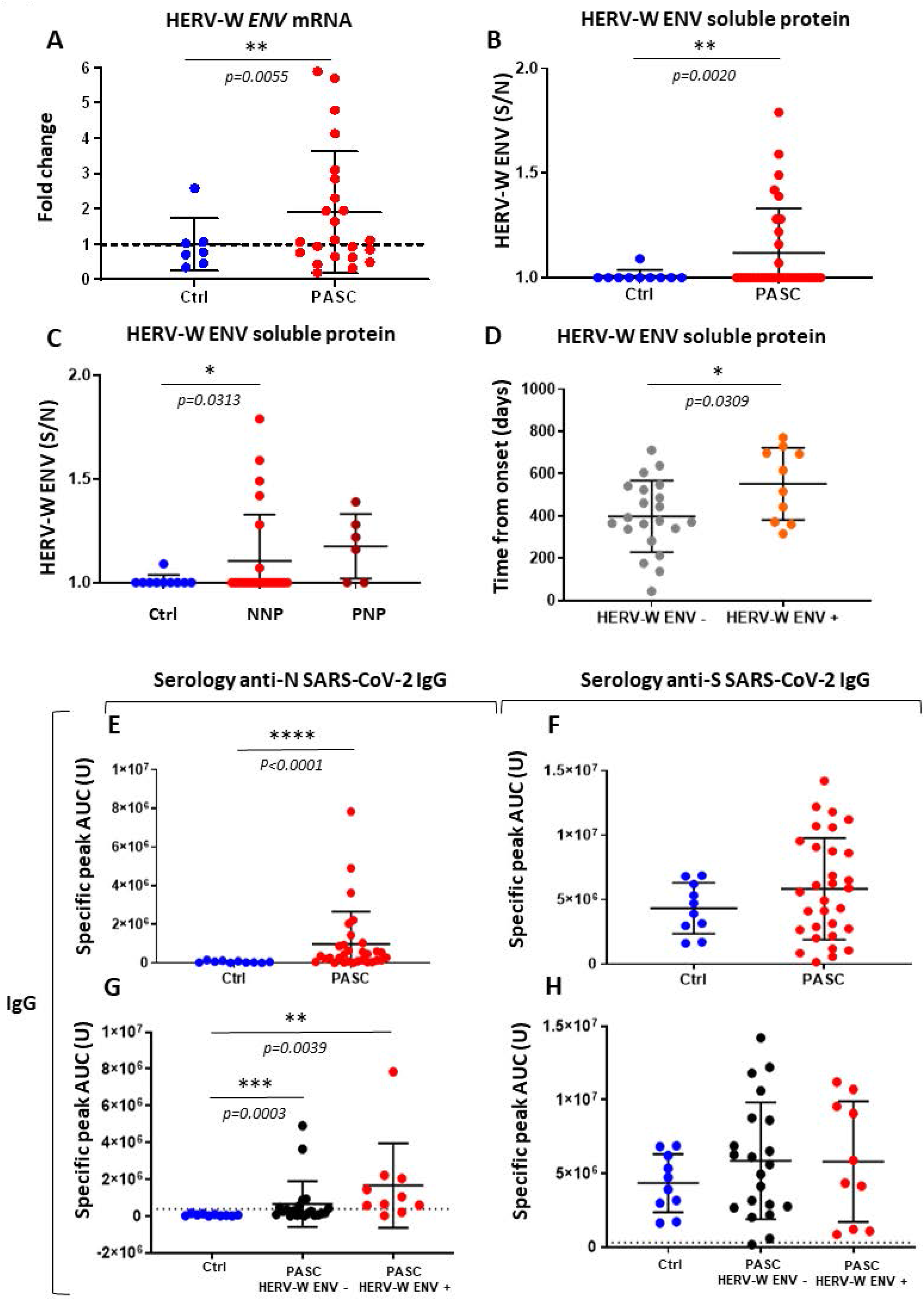
HERV-W ENV and Anti-Nucleocapsid IgG antibody are significantly detected in PASC. A, HERV-W *ENV* mRNA detection in PBMCs based on change fold of 2^-ΔΔCT^ compared to the mean of control groups; blue dots: controls group; red dots: PASC group. Here, a lower number of samples was analyzed, since 8 PASC and 3 controls yielded too low RNA concentration or did not pass QC (RIN<8). B, HERV-W ENV soluble oligomeric protein detection in serum of patients, results are expressed on total signal normalized with the mean of background noise measured on control cohort; blue dots : controls group; red dots : PASC group. C, post-hospitalized subgroup (PNP) is extracted from the PASC group and compared to PASC subgroup not hospitalized during acute COVID-19 (NNP); blue dots: controls group; red dots: NNP subgroup; dark red dots: PNP sub group. D, Comparison of the time from onset between HERV-W ENV- and HERV-W ENV+ PASC subgroups; grey dots: PASC HERV-W ENV-subgroup; orange dots: PASC HERV-W ENV+ subgroup. E, IgG anti-SARS-CoV-2 Nucleocapsid; F, IgG anti-SARS-CoV-2 Spike. Comparison between PASC HERV-W ENV- and PASC HERV-W ENV+ subgroups for IgG anti-Nucleocapsid (G), and anti-Spike (H). The value on the Y-axis (U) represents the area under the curve (AUC) of the electrophoregram specific peak corresponding to the immunodetected Ig isotype from tested sera against each antigen.

In Figure 1C PASC patients were stratified based on hospitalization status during acute SARS-CoV-2 infection. Positive cases for W-ENV protein represented about one fourth in the NNP subgroup (24%, 6/25), but a majority in the PNP subgroup (67%, 4/6). The difference of W-ENV signal to noise values (S/N) between NNP and controls was significant (Wilcoxon Signed Rank test: Ctrl W=1.00, PASC W=21.00, N=10+25, p=0.0313) but, probably due to the low numbers, not between PNP and Ctrl. Interestingly, W-ENV positive (W-ENV+) PASC patients tended to be further from the date of infection (Fig. 1D). A significant difference in the time from onset of acute COVID-19 between W-ENV+ and W-ENV-patients was confirmed (Unpaired t-test with Welch’s correction: t=2,346, df=17,61, N=21+10, p=0.0309). Some of these cases had suffered from post-COVID symptoms for two years after the acute infection, which corresponded to the beginning of the pandemic.

### SARS-CoV-2 -specific antibody responses

To study the humoral immune response to SARS-CoV-2 infection in PASC patients, we measured the levels of all immunoglobulins (IgG, IgM, IgA and IgE) versus the control group (including vaccinated individuals) against SARS-CoV-2 nucleocapsid (N) and spike (S) antigens.

Anti-N IgG were detected in a subgroup of PASC patients but not in controls (Wilcoxon Signed Rank test: Ctrl W=0.00, PASC W=444.0, N=10+31, p<0.0001; Fig. 1E), whereas no difference with Ctrl was observed for the anti-S IgG (Fig. 1F), which mostly indicated that Ctrl had been vaccinated with this antigen. Comparing W-ENV positive and negative PASC patients showed that anti-N IgG antibodies were detected in both subgroups, each one remaining significantly different from Ctrl (Wilcoxon Signed Rank test: Ctrl W=0.00, PASC/W-ENV-W=193.0, N=10+21, p=0.0003; Ctrl W=0.00, PASC/W-ENV+ W=53.0, N=10+10, p=0.0039; Fig. 1G), but no difference was seen between PASC subgroups and Ctrl for anti-spike IgG (Fig. 1H).

As shown in Figure 2, IgM against both S and N proteins were only observed in a subgroup of PASC patients with a wide range of titers. But, despite a few IgM positive PASC cases, a significant difference was seen versus controls for anti-N IgM (Wilcoxon Signed Rank test: Ctrl W=1600, PASC W=476.0, N=10+31 p=0.0480; Fig. 2 A) and, with high significance, for anti-S IgM (Wilcoxon Signed Rank test: Ctrl W=-6.00, PASC W=183.0, N=10+29, p<0.0001; Fig. 2 B). Of note, two sera with non interpretable technical profiles were omitted.. IgA antibodies levels were shown to be significantly elevated in PASC versus Ctrl for both N and S antigens, but anti-N IgA better discriminated PASC versus Ctrl (Wilcoxon Signed Rank test: anti-N IgA, Ctrl W=16.00, PASC W=476.0, N=10+31 p<0.0001; anti-S IgA, Ctrl W=-2.00, PASC W=273.0, N=10+31 p=0.0063; Fig. 2 C-D). Surprisingly, a subgroup of PASC patients had elevated IgE immunoglobulins against both N and S antigens (Wilcoxon Signed Rank test: anti-N IgE, Ctrl W=5.00, PASC W=336.0, N=10+31 p=0.0006; anti-S IgE, Ctrl W=14.00, PASC W=456.0, N=10+31 p<0.0001), with a large range of positivity versus all negative controls (Fig. 2 E-F).

**Figure 2.**
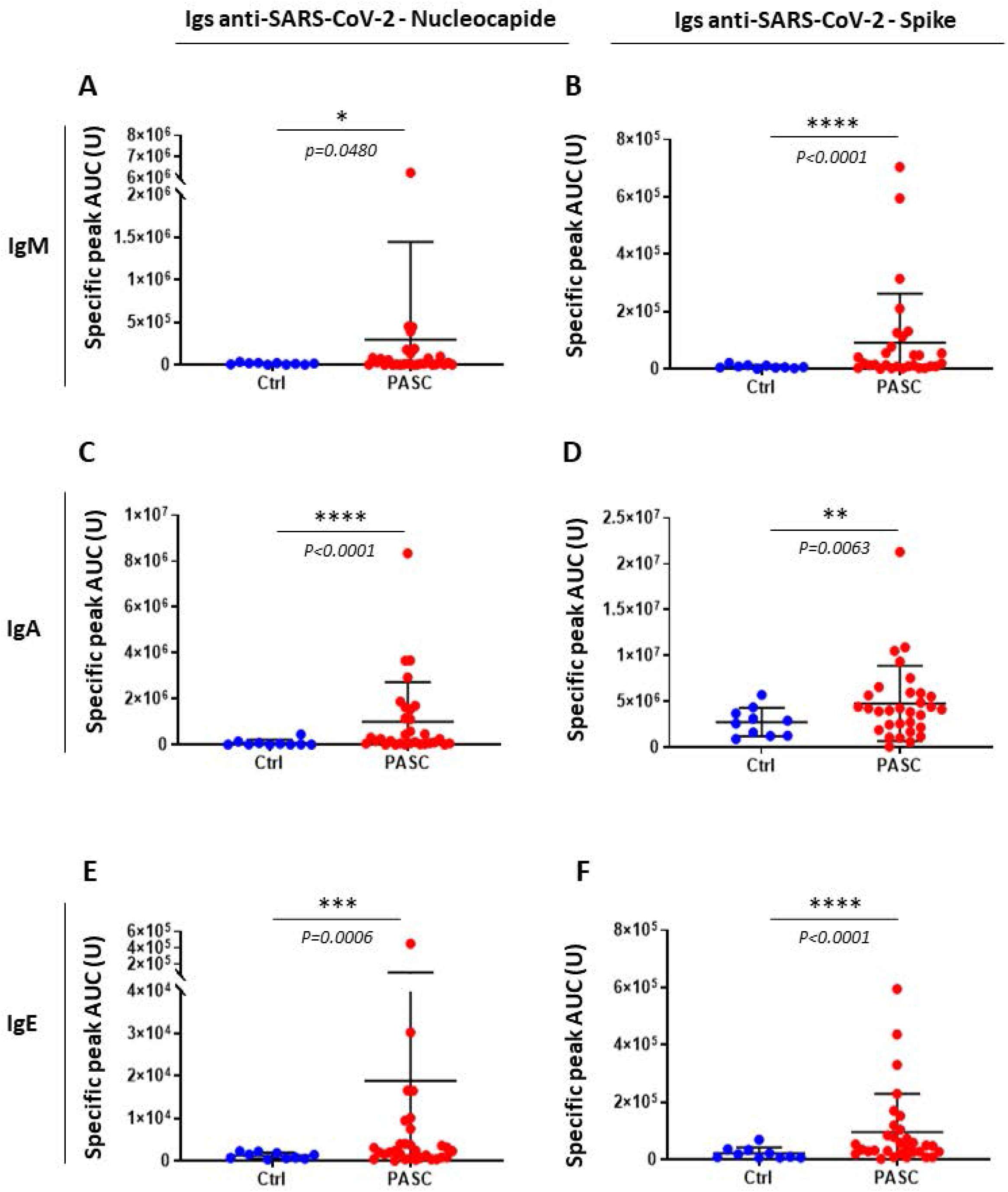
PASC serological IgM, IgA and IgE. A, IgM anti-SARS-CoV-2 Nucleocapsid; B, IgM anti-SARS-CoV-2 Spike IgM anti-S and N of two PASC patients with inconsistent results and diverging triplicate values were not be interpretable and therefore not taken into account in this analysis.; C, IgA anti-SARS-CoV-2 Nucleocapsid; D, IgA anti-SARS-CoV-2 Spike; E, IgE anti-SARS-CoV-2 Nucleocapsid and F, IgE anti-SARS-CoV-2 Spike. The value on Y axis (U) represents the area under the curve (AUC) of the electrophoregram specific peak corresponding to the immunodetected Ig isotype from tested sera against each antigen.

### IL-6 quantification

IL-6 was suggested to play a role in acute COVID-19 severity. In PASC, only a subgroup had elevated IL-6 (about 35%, 11/31) but, whereas most values were below the normal threshold (3pg/ml) in NNP, nearly all PNP (5/6) had elevated IL-6 levels in the serum with a significant difference from Ctrl (Mann Whitney test: U=3, N=10+6, p=0.0017;Fig. 3A). When W-ENV positive and negative PASC subgroups were compared, the W-ENV+ patients showed significantly higher IL-6 levels (Mann Whitney test: U=31, N=21+10, p=0.0011; Fig. 3B), consistently with the significant correlation found between PASC serum levels of HERV-W ENV and IL-6 (Spearman correlation test: r=0.5877, 95% CI 0.2847 to 0.7840, pairs N=31, p=0.0005; Fig. 3C). In figure 3D, PASC were stratified according to both anti-N SARS-CoV-2 IgG and W-ENV serum levels. All samples from the W-ENV+ subgroup had elevated IL-6 levels, when compared to all other W-ENV negative sera with or without anti-N IgG (Mann Whitney test: W-ENV+/IgG N+ Vs W-ENV-/IgG N-, U=7, N=8+9, p=0.0035; W-ENV+/IgG N+ Vs W-ENV-/IgG N+, U=10, N=8+13, p=0.0013; Fig. 3D).

**Figure 3.**
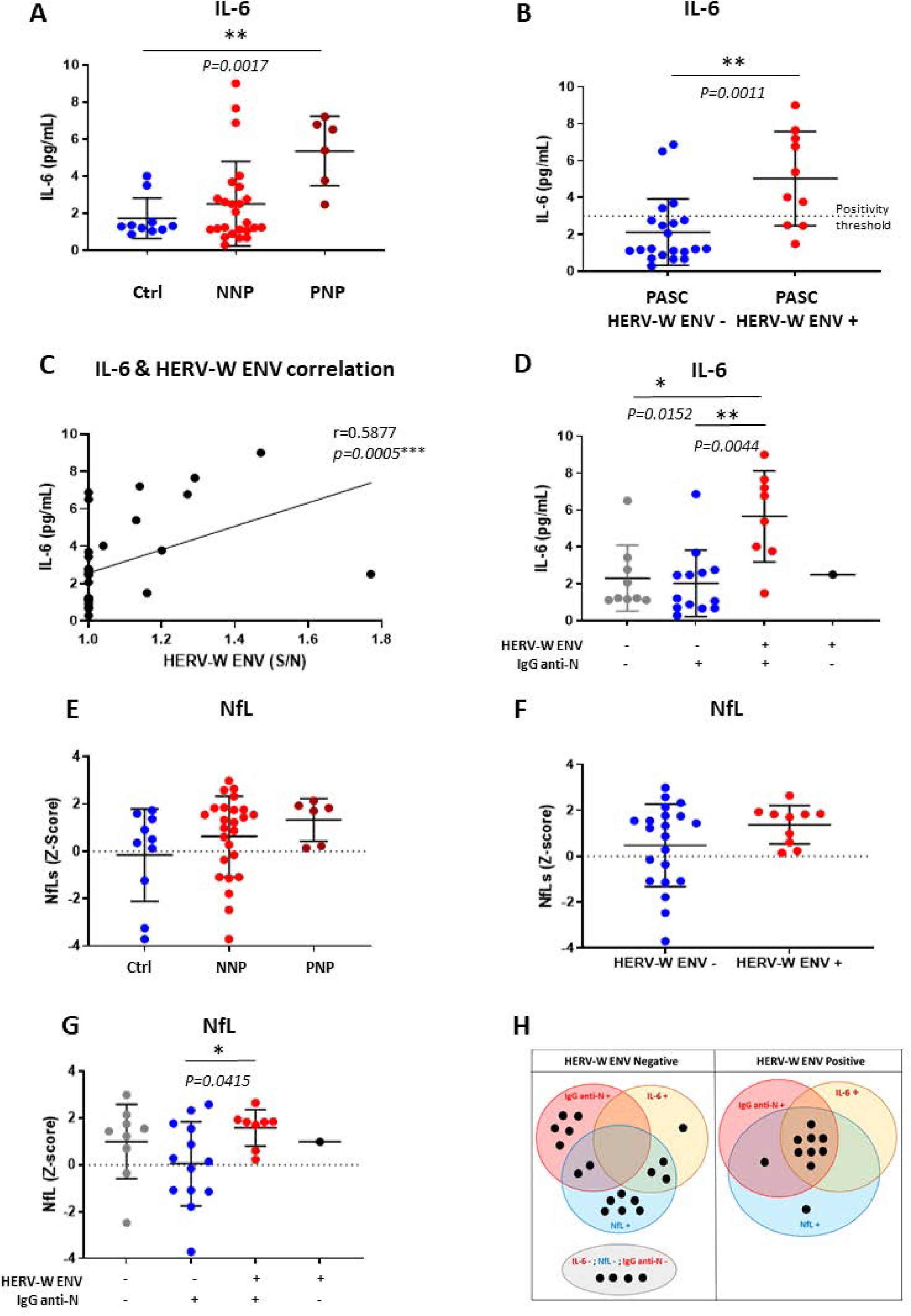
Interleukine-6 (IL-6), neurofilament light chain (NfL) biomarkers and combination with HERV-W ENV or anti-nucleocapsid IgG (IgG anti-N) status stratifying PASC patients. A, IL-6 dosages for controls (bleu dots), NNP (red dots) and PNP (dark red dots). B, IL-6 dosage comparisons between HERV-W ENV- and HERV-W ENV+ subgroups in PASC cohort. C, correlation between IL-6 and HERV-W ENV levels in serum. D, IL-6 levels in PASC patients stratified by: HERV-W ENV- / IgG anti-N- (grey dots), HERV-W ENV- / IgG anti-N+ (blue dots) and HERV-W ENV+ / IgG anti-N+ (red dots). E, NfL Z-Scores of controls (blue dots), NNP (red dots) and PNP (dark red dots). NfL Z-scores comparisons between HERV-W ENV- and HERV-W ENV+ subgroups in PASC cohort. G, NfL levels in different PASC subgroups: HERV-W ENV- / IgG anti-N- (grey dots), HERV-W ENV- / IgG anti-N+ (blue dots) and HERV-W ENV+ / IgG anti-N+ (red dots). H, Graphical presentation of patients’ clusters defined by the selected serum biomarkers, within HERV-W ENV negative and HERV-W ENV positive subgroups. The sera positive for each biomarker are represented by dots in the colored circles; those negative for all parameters are in the grey oval.

### Neuronal impairment biomarker: Neurofilament Light chain (NfL)

We measured the levels of serum NfL in PASC with cognitive impairment, since shown to be an accurate peripheral biomarker of neuronal degeneration or injury within the central nervous system^27^ (Figure 3E-G). Results were normalized with Z-score according to the age of the patients or control individuals, as already defined ^28^.

No significant difference was found between the distributions of NfL serum levels in PNP versus NNP, nor Vs Ctrl (Fig. 3 E). Nonetheless, when stratified with W-ENV, all (100%, 10/10) W-ENV+ patients had elevated levels of serum NfL while only 62% (13 out of 21) of W-ENV-negative sera had a NFL Z-scores above 0 (Fisher exact test = 0.0317 p<0.05, α=1, N=31; Fig. 3 F). Moreover, Z-scores were all above zero in the subgroup with both W-ENV and anti-N IgG positivity which was significantly different from the anti-N IgG+ and W-ENV-subgroup (Unpaired t test with Welch’s correction: t=2,688, df=17,61, N=22, p= 0,0152; Fig. 3G),.

### Biomarker-defined subgroups

Figure 3H represents clusters of all PASC patients studied for the above-mentioned serum biomarkers. Within the W-ENV+ subgroup the majority (7/10) of PASC were positive altogether for NfL, IL-6 and IgG anti N-SARS-CoV-2 but only one (1/21) in the W-ENV negative subgroup (Chi-square statistic=17.890, *p*=0.00002, α=1, N=31 tested on all parameters). All W-ENV+ Neuro-PASC had positive serum levels of NfL (10/10). Moreover, within the W-ENV + subgroup, the NfL+ cluster included all IL-6+ or Ig anti-N+ sera and therefore coincided with W-ENV positivity in Neuro-PASC patients, i.e., defined the same subgroup when combined with W-ENV positivity.

Matching with acute COVID effective hospitalization also, all HERV-W ENV+ patients from the PNP subgroup were positive for the three other biomarkers (NfL, IL-6 and IgG anti-N; data not shown).

Within the W-ENV-subgroup, a single case was positive for NfL, IL-6 and IgG anti-N altogether. As found to be the core parameters in the W-ENV+ subgroup, this may question the sensitivity of the present test for HERV-W ENV soluble antigen detection in serum. The patterns for single or double positivity appeared to be variable while only NfLs positive levels were dominant, thereby suggesting relationship with the neuro-cognitive impairment in this cohort. However, a cluster was clearly identified to be negative for all these biomarkers, beyond negativity for W-ENV as well. This “all negative” cluster is consistent with very different pathogenic factors involved in their post-COVID symptomatology.

Of note, though all W-ENV+ patients had positive Z-scores for NfL, taken alone this biomarker revealed dominant in all Neuro-PASC cases, W-ENV+ and W-ENV- (Figure 3F). Negative Z-scores were found in few cases positive for IL-6 only or for IgG anti-N only but, most interestingly, within the isolated cluster negative for all tested biomarkers (figure 3H).

SARS-CoV-2 serology showed significant differences with controls for anti-N IgG (Figure 1E) and for anti-N and/or anti-S IgM, IgA or IgE (Figures 2A-F). But, even in combination with other biomarkers, it did not provide a clear-cut stratification when considered without the other studied biomarkers.

Therefore, Neuro-PASC subgroups could be defined as (i) W-ENV+ and NfL+, (ii) W-ENV- and (NfL+ or IL-6+ or IgG anti-N+) and (iii) negative for all the tested parameters. In addition, possible indication may be also provided on SARS-CoV-2 persistence by IgM, on the route of infection in extrapulmonary cases^29^ or on the site of persistence by IgA and, particularly, on potentially associated immunoallergic symptoms by IgE.

Finally, using data from the PROMIS scale^30^ and other clinical data showed no significant difference in the distribution of results between the subgroups consistently defined by the different biomarkers as exemplified with HERV-W ENV and anti-N-SARS-CoV-2 IgG in Figure 4.

**Figure 4.**
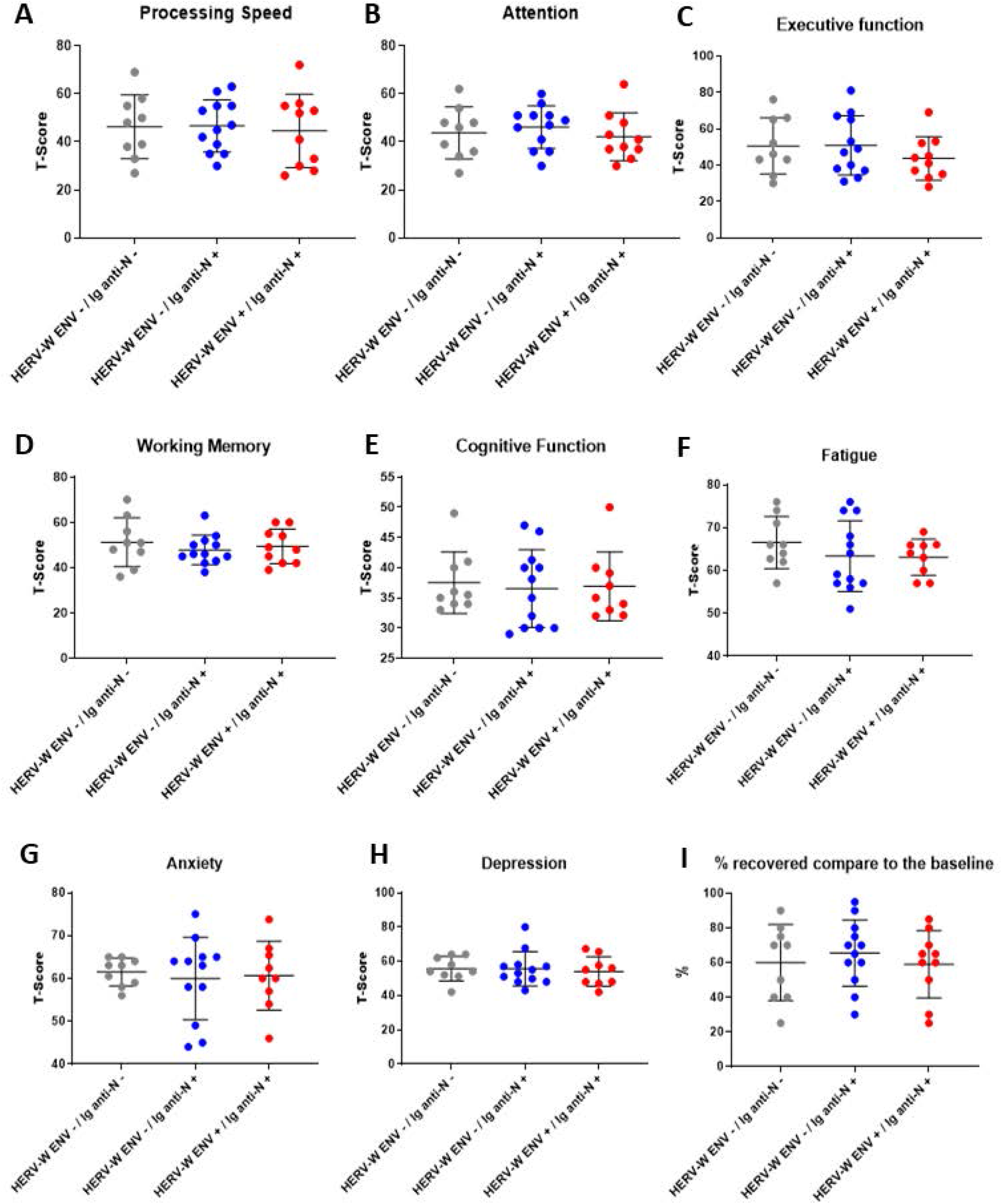
Comparison of clinical data between groups based on HERV-W ENV and serology profiles. Three groups from the PASC cohort are represented based on main biomarkers results: HERV-W ENV+ /Igs anti-SARS-CoV-2 Nucleocapsid (Ig anti-N) + ; HERV-W ENV- /Igs anti-SARS-CoV-2 Nucleocapsid (Ig anti-N) + and HERV-W ENV- /Igs anti-SARS-CoV-2 Nucleocapsid (Ig anti-N). Several parameters were studied: A, Processing speed; B, Attention; C, Executive function; D, Working Memory; E, Cognitive function; F, Fatigue; G, Anxiety; H, Depression and I, Recovery compared to the baseline.

### Conclusion

The present results, despite the limited number of patients, consistently identified three major Neuro-PASC subgroups with significant differences stratified according to the presence or absence of the HERV-W ENV protein combined with positive Z-score for NfLs in serum. SARS-CoV-2 serology with all Ig isotypes should also provide useful indications on the infectious status, with possible viral persistence or a possible immunoallergic specific phenotype linked to elevated anti-spike or Nucleocapsid IgE. Since clinical data could not differentiate consistent subgroups of patients, unlike biological parameters, the biomarker-defined subgroups or clusters may be considered for differing therapeutic indications in post-COVID treatment. This should be a prerequisite for clinical trials to avoid mixing patients with and without the pathogenic target of a given drug, which may generate non-significant results versus placebo, despite possibly responding cases. Thus, the presented biomarker and serology-based stratification should pave the way to precision medicine in post-acute COVID sequalae or syndromes. Based on the present proof of concept and on that of other studies, a diagnostic-based therapy with an anti-HERV-W ENV neutralizing antibody (Temelimab) is being evaluated in a clinical trial with PASC patients positive for the HERV-W ENV protein in serum (ClinicalTrials.gov Identifier: NCT05497089).

## Data Availability

All data produced in the present study are available upon reasonable request to the authors

## Author Contributions

IK : conceptualization of the study, evaluating all patients in the Neuro-COVID-19 clinic, supervising patients sample collection, manuscript editing and review

NW collaborators: Zack and Millenia: organized sample and data collection from all Neuro-PASC and control patients at Northwestern University, manuscript review

Barbara and Lavanya: Conceptualization of the study, assisted in sample and data collection from all Neuro-PASC and control patients at Northwestern University, editing and reviewing the manuscript

HP: conceptualization, supervision, formal analysis, investigation, data curation, writing and manuscript review.

AL, CB, SF: HERV-W-ENV detection, NfL, cytokines HERV-W-ENV detection, NfL, cytokines, serological analyses.

B.C.: methodology, management interpretation of analyses and manuscript review.

J.P: Serological and antigenemia analyses; graphs, figures drawing and manuscript review.

J.B.: PCR, serological and antigenemia analyses; graphs, figures drawing and manuscript review.

H.P. and E.O.: conceptualization, funding acquisition, supervision, formal analysis, investigation, data curation, writing original draft and manuscript review.

All authors contributed to the article and approved the submitted version.

